# Characteristics and Circumstances of US Overdose Deaths Identified as Heat-Related

**DOI:** 10.64898/2026.05.11.26352941

**Authors:** Manuel Cano, Chung Jung Mun, Kaylin Sweeney, Raminta Daniulaityte

**Author notes:** **Corresponding author:** Manuel Cano, 411 N Central Ave Suite 863, Phoenix, AZ 85004, USA. **Sources of support/funding:** Funding was received from the Arizona State University College of Health Solutions Heat and Health seed grant (PI Daniulaityte). Conflicts of interest: None.

## Abstract

**Objectives:** To examine the extent to which heat-related causes of death are recorded in fatal drug overdoses, how these patterns vary across states and over time, and how overdose characteristics differ between deaths with, versus without, heat involvement recorded.

**Methods:** Death certificate data for all drug overdose deaths in US residents from 2001 to 2024 (from the National Center for Health Statistics) were analyzed to identify whether a heat-related cause of death was also listed on the death certificate. Joinpoint regression, descriptive statistics, and nonparametric tests were used to examine temporal trends and compare overdose deaths with versus without recorded heat involvement.

**Results:** In 2001, fewer than 10 drug overdose deaths with recorded heat involvement were identified, but this number increased to 558 in 2024. From 2013 to 2024, mortality rates increased significantly, with an estimated annual percent change of 30.1 (95% Confidence Interval, 26.5–47.1). The highest mortality rates and numbers of deaths were observed in residents of Arizona and Nevada. American Indian/Alaska Native, Mexican-heritage, and foreign-born populations accounted for larger shares of overdose deaths with, compared to without, heat involvement recorded. A street or highway was more frequently identified as the place of injury in overdose deaths with (18.9%), versus without (2.2%) heat involvement reported. Psychostimulants such as methamphetamine were involved in 85.9% of overdose deaths with, compared to 28.9% without, recorded heat involvement.

**Conclusions:** Although representing only a fraction of all overdose deaths, fatal overdoses involving heat exposure have increased markedly over time and disproportionately impact certain states and demographic groups.

## Background

### Characteristics and Circumstances of US Overdose Deaths Identified as Heat-Related

Exposure to extreme heat is associated with numerous health risks (Ebi et al., 2021), but relatively less is known about the potential role of heat in drug overdose deaths. Drug overdoses represent a leading cause of injury-related death in the United States (US) and in several other nations (Martins et al., 2015). As environmental heat is projected to continue rising (Lizana et al., 2026), a better understanding of heat involvement in drug overdose deaths may become an increasingly important component of comprehensive overdose prevention efforts in the years ahead.

Several studies have documented a relationship between environmental heat and overdoses. For example, four different analyses of temperature data and overdose deaths in New York City, Quebec, and British Columbia found that higher daily or weekly temperatures were associated with more fatal overdoses involving cocaine (Auger et al., 2017; Bohnert et al., 2010; Henderson et al., 2023; Marzuk et al., 1998). In these studies, the association with heat was limited to cocaine-involved overdose deaths. Nonetheless, in an analysis of drug-related emergency department (ED) visits in California, higher ambient temperature was positively associated with ED visits for overdoses involving several different drug types, including amphetamines, cocaine, and opioids (Chang et al., 2023).

In a more recent study of counties in the contiguous US, significant associations between county-level heat index and fatal overdoses were observed across deaths involving opioids, cocaine, and psychostimulants such as methamphetamine (Dennett et al., 2026). This analysis estimated that approximately 3,310 excess drug overdose deaths over the period of June-September 1999-2020 could be attributable to heat exposure in the contiguous US (Dennett et al., 2025). Like many other studies, this work employed weather data to estimate the role of heat in overdose deaths. A complementary approach is to directly examine overdose deaths in which heat was recorded as a contributing cause of death on the death certificate. Using this approach, an analysis of data from the State Unintentional Drug Overdose Reporting System (SUDORS) in Arizona examined deaths in which both drug overdose and heat-related causes were recorded on the death certificate, integrating data from medical examiner reports, postmortem investigations, and toxicology testing (Cano et al., 2025). Results indicated that in Arizona, heat involvement was recorded in 3% of drug overdose death certificates in 2019 and 15% in 2023, and methamphetamine was detected in 92% of overdose deaths with documented heat involvement (Cano et al., 2025). What remains unclear is the extent to which some of these findings from Arizona, one of the hottest US states (with temperatures reaching or exceeding 110 degrees each July in the Phoenix area; National Weather Service, 2026), generalize to other areas in the US.

To fill this gap in the literature, we examined individual-level data from drug overdose deaths across the US that specifically list heat or hyperthermia as a contributing cause of death on the death certificate. Whereas prior research used heat index data to statistically estimate the number of drug overdose deaths attributable to heat in the US (Dennett et al., 2026), the present study directly quantifies how many death certificates for drug overdoses mention heat or hyperthermia and how this varies by state and has changed over time. In addition, this study compares overdose deaths with versus without documented heat involvement, focusing on basic demographics of decedents as well as additional decedent-level factors that prior literature has highlighted as particularly salient for heat exposure or heat-related deaths, such as nativity (Moyce & Schenker, 2017; Taylor et al., 2018) and occupation (Petitti et al., 2013). Finally, the study examines several contextual features of overdose deaths with recorded heat involvement, including place of injury and death, as well as the specific types of drugs involved, informed by prior literature indicating how stimulant drugs such as methamphetamine can increase body temperature and impair thermoregulation (Matsumoto et al., 2014).

## Methods

This analysis of de-identified secondary data was classified by the Arizona State University Institutional Review Board as “not research involving human subjects.”

### Data Source and Sample

Data were drawn from the 2001-2024 restricted-access National Center for Health Statistics (NCHS) Multiple Cause of Death files (National Center for Health Statistics, 2026), which compile death certificate records for all US resident deaths across the 50 states and the District of Columbia. In this dataset, each death is coded with one “underlying cause of death” and up to 20 additional “multiple causes of death.” Our analytic sample of drug overdose deaths (of any intent and involving any drug) was identified via an “underlying cause of death” coded with any International Classification of Diseases, Tenth Revision (ICD-10) code of X40-44 (accidental drug poisoning), X60-64 (intentional drug self-poisoning), X85 (drug poisoning classified as homicide), or Y10-14 (drug poisoning of undetermined intent). Population estimates (for calculating mortality rates) were obtained from the CDC WONDER online platform (Centers for Disease Control and Prevention, 2026), which provides US Census Bureau estimates of the resident population.

### Measures

The primary measure of interest was whether heat/hyperthermia was recorded as a contributing cause of death on the death certificate. On death certificates, causes of death are reported by a medical certifier such as a medical examiner or coroner. We defined heat involvement via the presence of any “multiple cause of death” corresponding to ICD-10 code X30 (“exposure to excessive natural heat-hyperthermia”) or T67 (“effects of heat and light,” spanning T67.0-T67.9), and we excluded any death coded with W92 (“exposure to excessive heat of man-made origin”), consistent with methodology used in publications of the US Environmental Protection Agency (EPA, 2017).

Additional measures of interest comprised characteristics of the decedent and the overdose death. Decedent characteristics included demographics: age (in years); sex (male or female, as recorded on the death certificate); educational attainment (in categories of 8th grade or less, 9-12th grade with no diploma, high school graduate or General Educational Development [GED], some college but no degree, associate or bachelor’s degree, and graduate degree); and marital status (categorized as single/never married, married, divorced, and widowed). We also assessed race/ethnicity, using the most detailed categories available that included at least 10 overdose deaths with heat recorded over the analytic period. Racial categories with low numbers of deaths were combined into broader categories as needed to reach the threshold of 10, resulting in the following mutually-exclusive categories: Non-Hispanic (NH) White; NH Black; NH American Indian/Alaska Native (AI/AN); NH Asian/Pacific Islander (combined due to low numbers); NH Black and White; NH AI/AN and White; NH Other Multiple Race; Mexican; Puerto Rican; and other Hispanic. Nativity (coded as born in the 50 states/DC/territories versus foreign-born) was also included as a measure of interest based on prior studies indicating that immigrants to the US are disproportionately employed in outdoor jobs (e.g., agriculture, construction, landscaping; Moyce & Schenker, 2017), and that non-citizens experience increased risk of heat-related mortality in the US (Taylor et al., 2018). Usual occupation during lifetime was also examined, using binary (yes/no) indicators for whether the death certificate indicated that the decedent’s usual occupation (regardless of whether they were still employed at the time of death) fell into any of three different categories associated with environmental heat exposure or heat-related injury/death in prior literature (Gubernot et al., 2015; Kearney & Imai, 2023; Petitti et al., 2013): (1) construction/extraction; (2) building/grounds cleaning/maintenance; and (3) farming/fishing/forestry.

Circumstances/characteristics of the overdose death included: the year and month of death; the state and US Census Division of the decedent’s residence; whether the death certificate indicated that the injury (overdose) occurred at work (yes/no); whether the reported place of injury was at home (yes/no); whether the reported place of injury was on the street/highway (yes/no); the place of death (medical center, home, hospice/nursing facility/long-term-care, or “other”); and the drugs involved. Drug categories were based on ICD-10 codes from contributing causes of death, comprising: synthetic opioids such as fentanyl (code T40.4 for “synthetic opioids excluding methadone”); psychostimulants such as methamphetamine (code T43.6 for “psychostimulants with abuse potential, excluding cocaine”); and cocaine (code T40.5 for “cocaine”). These three drug types were selected because they represent the drugs most frequently reported in overdose deaths in the US (Garnett et al., 2026); we also examined common combinations of these drug classes (i.e., psychostimulants plus synthetic opioids, cocaine plus synthetic opioids).

### Statistical Analyses

Analyses (and data visualizations) were conducted in Stata/MP 19.5, Joinpoint Trend Analysis Software version 6.0.1 (Surveillance Research Program, National Cancer Institute, 2026), and Python 3.12.7. All analyses used listwise deletion for missing data; supplemental Table S1 provides details on the extent of missing data observed for each measure used in the study. First, we calculated US mortality rates by *year* (2001-2024) for overdose deaths with heat-involvement recorded on the death certificate. Mortality rates were expressed per 100,000 (representing crude rates, calculated as the number of deaths divided by the population size and then multiplied by 100,000). We used Joinpoint regression (Kim et al., 2000) to examine trends from 2005-2024, with 2005 the first year included because heat-related overdose deaths in several previous years did not reach 10 deaths per year (the threshold for presenting data, per NCHS). We specified a relatively conservative Joinpoint model with first-order correlated standard errors in a log-linear specification that calculates annual percent change (APC).

Next, we calculated mortality rates per 100,000 for each *state*, combining data from the entire study period (2001-2024) for more robust estimates. “State” was defined as the state of the decedent’s residence. We used a choropleth map to depict state-level rates; rates based on fewer than 10 deaths were not presented, in compliance with NCHS privacy regulations.

To visualize differences by month of death, we used a bar plot to depict the percentage of all overdose deaths (2018-2024 combined) that mentioned heat as a contributing cause of death, for each month individually. Finally, we examined decedent characteristics and overdose circumstances for: a) overdose deaths with heat-involvement recorded; and b) overdose deaths without heat involvement recorded. We compared the two groups using chi-square tests for categorical measures and a Wilcoxon rank-sum test for the one continuous measure. In this analysis, we focused on the seven most recent years (2018-2024) because changes over time in the coding of some demographic measures (e.g., race; Heron, 2021) rendered measures from earlier years not directly comparable. Occupation categories were examined only for years 2022-2024 because not all states reported this measure in prior years. Marital status, occupation, and whether the injury occurred at work were examined only in decedents aged 18 or older, and educational attainment was assessed only in decedents aged 25 or older.

### Supplemental Analysis

Considering that residents of one state, Arizona, accounted for 44% of all overdose deaths with heat involvement recorded during the study period, we conducted a supplemental analysis to assess whether study findings were driven by this state. After excluding deaths among Arizona residents, we repeated the descriptive analyses comparing characteristics of overdose deaths with (versus without) recorded heat involvement.

## Results

### Trends over Time

Drug overdose deaths with heat involvement recorded on the death certificate rose from fewer than 10 in 2001 to 558 in 2024. As depicted in Figure 1, results of Joinpoint regression analyses (examining rates from 2005-2024) indicated that mortality rates for overdoses with heat involvement recorded did not significantly increase from 2005-2013 but significantly increased from 2013-2024 (p<0.001), with an estimated annual percent change of 30.1 (95% Confidence Interval, 26.5–47.1).

**Figure 1.**
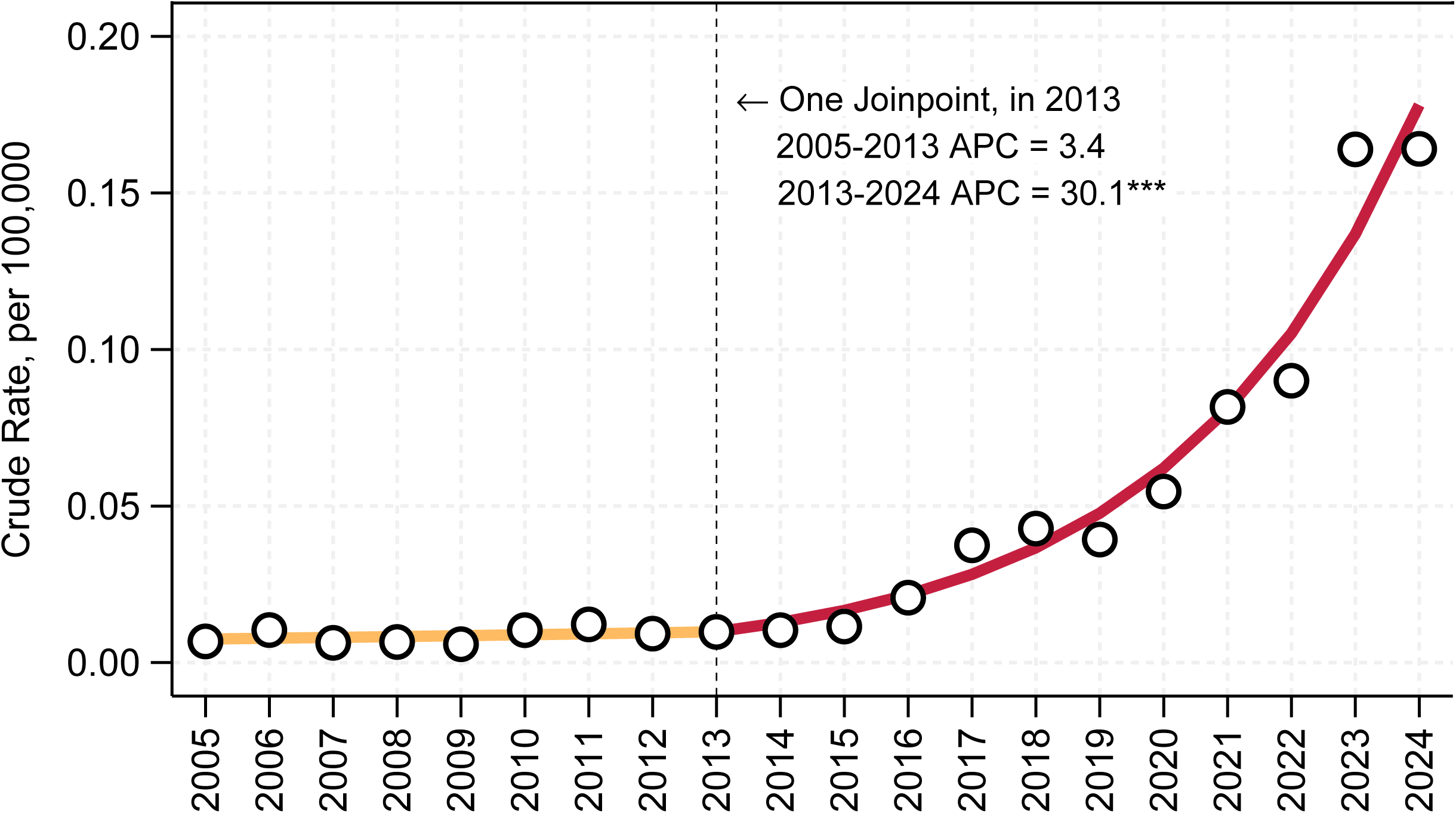
Trend in Mortality Rates for Drug Overdoses with Heat Involvement Recorded, United States, 2005-2024. *Notes*. Death counts from the restricted-access Multiple Cause of Death files from the National Center for Health Statistics; population estimates from the US Census Bureau via CDC WONDER. The markers (white circles with black borders) denote observed rates. The fitted line and Joinpoint estimates were calculated using Joinpoint Trend Analysis Software, Version 6.0.1. *** p < .001. *Abbreviations*: APC, annual percent change.

### State-Level Variation

Figure 2 depicts 2001-2024 state-level mortality rates for overdose deaths with heat involvement recorded; rates are not displayed for states with fewer than 10 deaths (in accordance with NCHS privacy regulations). The highest mortality rates were observed in residents of Arizona (0.75 per 100,000) and Nevada (0.66 per 100,000). As detailed in supplemental Table S2, the highest absolute numbers of overdose deaths with heat involvement recorded (2001-2024) were in residents of Arizona (1,185), Nevada (436), California (258), and Texas (180). Among states with at least 10 overdose deaths with heat recorded during the study period (2001-2024), the proportion of overdose deaths that also listed heat on the death certificate ranged from 0.02% in North Carolina and 0.03% in New York to 2.8% in Nevada and 3.5% in Arizona (supplemental Table S2).

**Figure 2.**
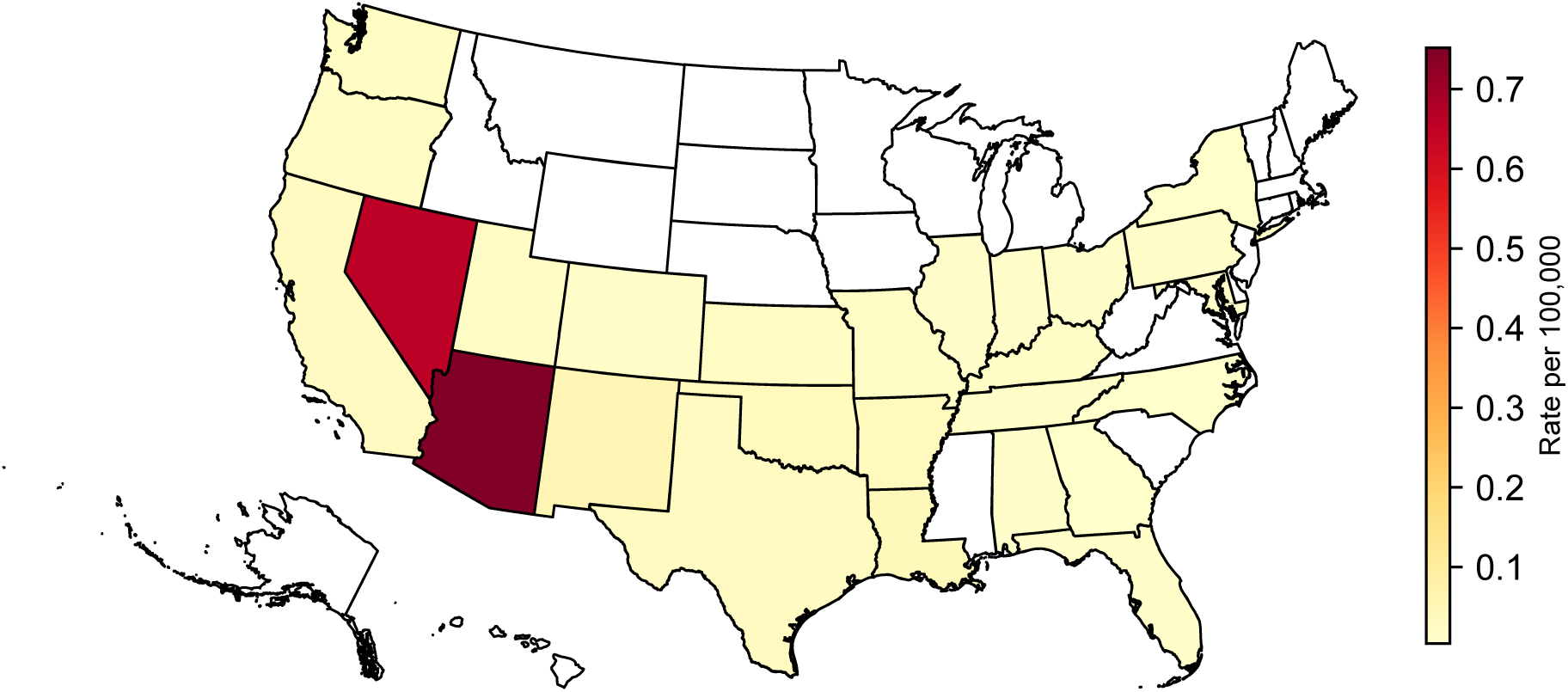
State-level Mortality Rates for Drug Overdoses with Heat Involvement Recorded, United States, 2001-2024. *Notes*. Death counts from the restricted-access Multiple Cause of Death files from the National Center for Health Statistics; population estimates from the US Census Bureau via CDC WONDER. Due to confidentiality restrictions, data are presented only for states with at least 10 drug overdose deaths involving heat over the study period.

### Characteristics of Overdose Deaths With vs. Without Heat Involvement Reported

As depicted in Figure 3, over the period of 2018-2024, heat involvement was recorded on the death certificate of 1.9% of all drug overdose deaths that occurred in the month of July, 0.9% of those that occurred in August, and 0.6% of those that occurred in June. Table 1 summarizes selected characteristics and circumstances of overdose deaths with and without heat involvement reported on the death certificate, also focusing on 2018-2024. With respect to decedent characteristics, individuals who died of an overdose with heat involvement recorded had a median age of 45, and individuals identified as female were underrepresented (15.3%) compared with their representation in overdoses without heat involvement recorded (30.7%). NH White Americans accounted for 53.5% of overdose deaths with, versus 66.1% of overdose deaths without, heat involvement recorded. NH American Indian/Alaska Native, Mexican heritage, and foreign-born populations each accounted for larger shares of overdose deaths with, compared to without, heat involvement recorded. Lower educational attainment levels were relatively more frequently reported in overdose deaths with, versus without, heat involvement reported (among decedents ages 25+). The share of decedents with a usual occupation of construction/extraction, building/grounds cleaning/maintenance, or farming/fishing/forestry did not significantly differ between overdose deaths with and without heat recorded. Finally, overdose deaths with heat recorded were concentrated in the Mountain Census Division, which accounted for 67.8% of these deaths versus 7.5% of deaths without heat involved.

**Figure 3.**
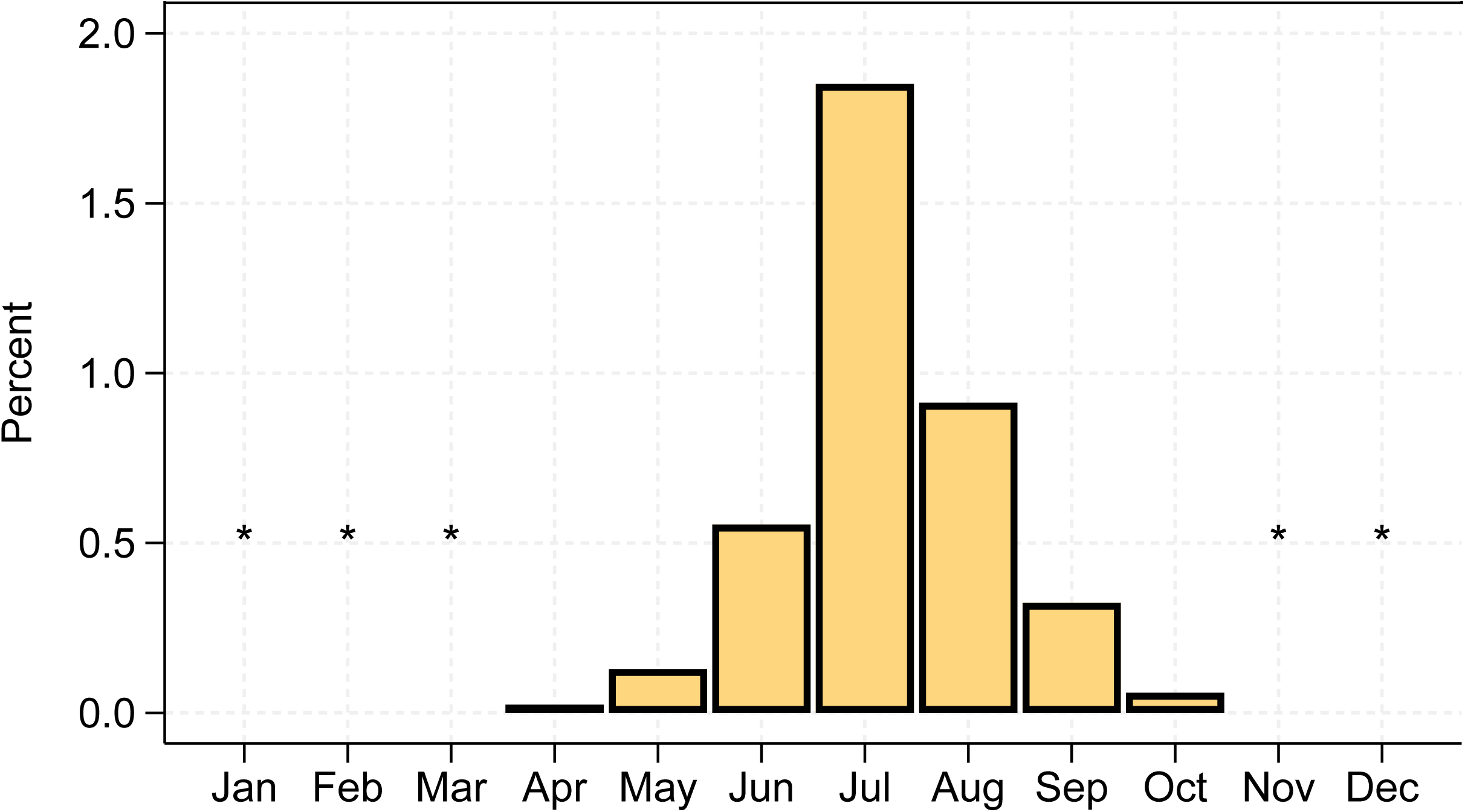
Drug Overdose Deaths with Heat Involvement Recorded, as a Percentage of All Overdose Deaths, United States, 2018-2024. *Notes*. A total of 628,827 overdose deaths occurred during the analysis period (2018-2024). Data from the restricted-access Multiple Cause of Death files from the National Center for Health Statistics. *Not presented if fewer than 10 overdose deaths with heat involvement were recorded in the given month (over the years 2018-2024 combined).

**Table 1.**
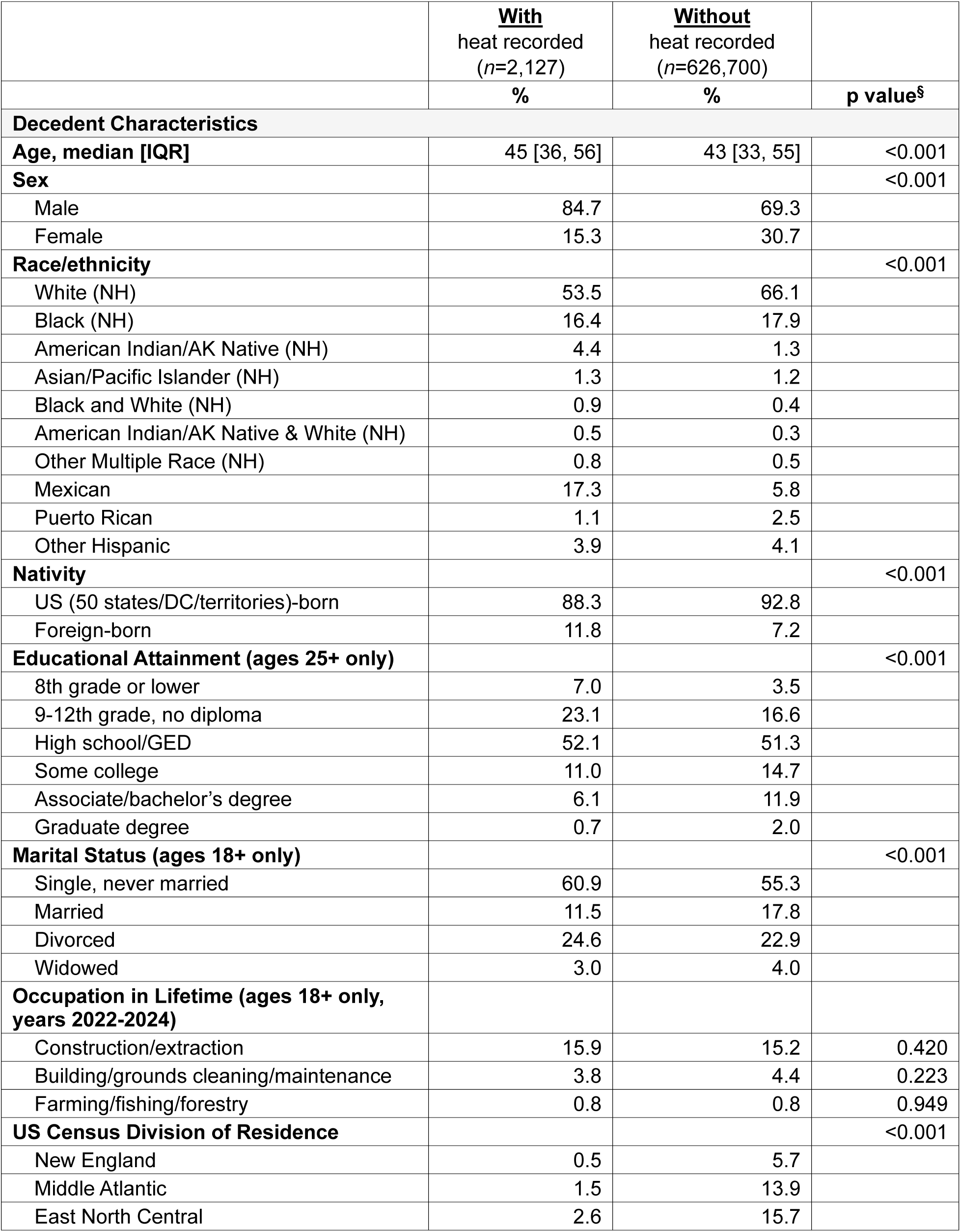
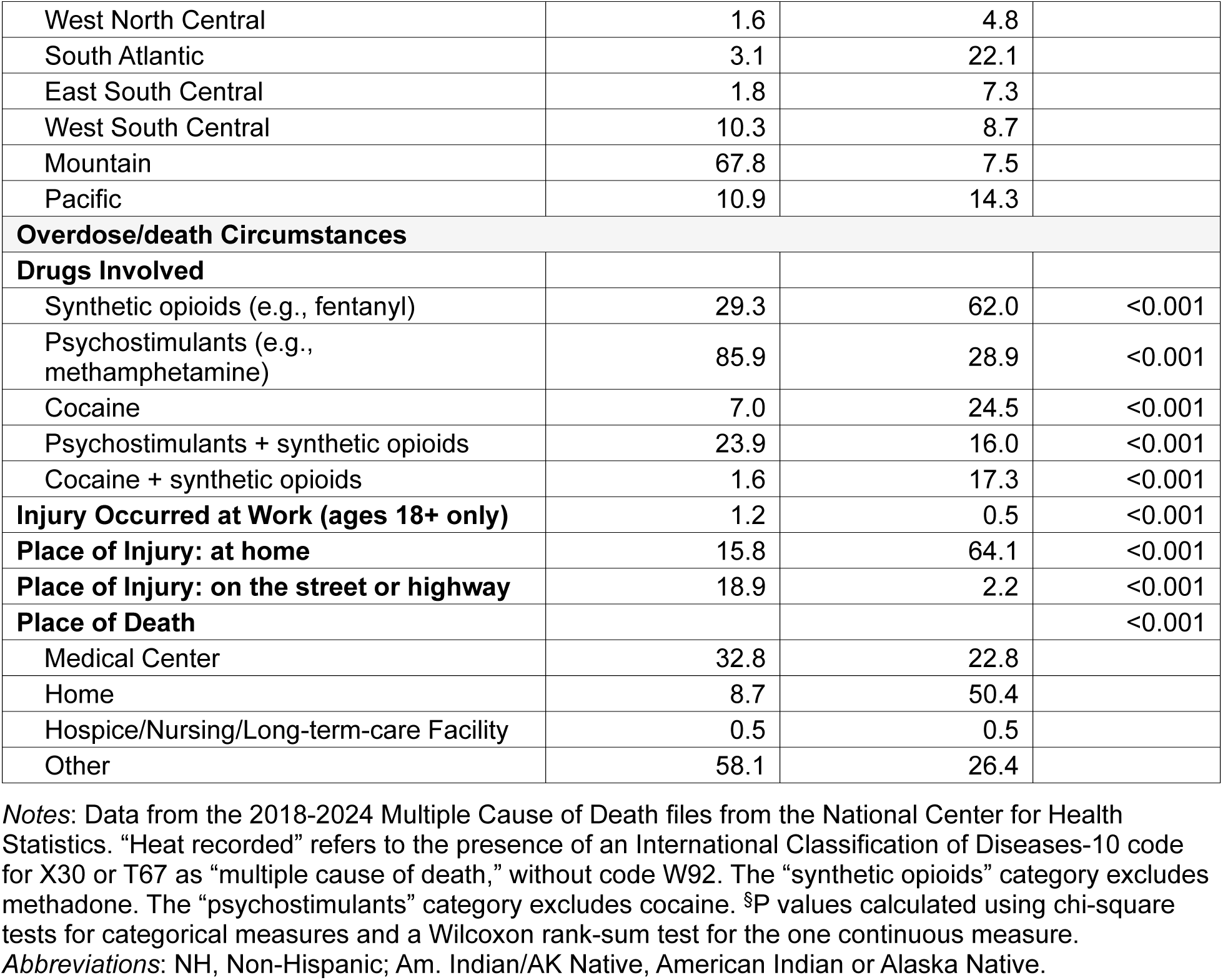
Characteristics and circumstances of overdose deaths with, versus without, heat involvement recorded, United States, 2018-2024.

|  | <u>With</u><br>heat recorded<br>(n=2,127) | <u>Without</u><br>heat recorded<br>(n=626,700) |  |
| --- | --- | --- | --- |
|  | % | % | p value<sup>s</sup> |
| **Decedent Characteristics** |  |  |  |
| **Age, median [IQR]** | 45 [36, 56] | 43 [33, 55] | <0.001 |
| **Sex** |  |  | <0.001 |
| Male | 84.7 | 69.3 |  |
| Female | 15.3 | 30.7 |  |
| **Race/ethnicity** |  |  | <0.001 |
| White (NH) | 53.5 | 66.1 |  |
| Black (NH) | 16.4 | 17.9 |  |
| American Indian/AK Native (NH) | 4.4 | 1.3 |  |
| Asian/Pacific Islander (NH) | 1.3 | 1.2 |  |
| Black and White (NH) | 0.9 | 0.4 |  |
| American Indian/AK Native & White (NH) | 0.5 | 0.3 |  |
| Other Multiple Race (NH) | 0.8 | 0.5 |  |
| Mexican | 17.3 | 5.8 |  |
| Puerto Rican | 1.1 | 2.5 |  |
| Other Hispanic | 3.9 | 4.1 |  |
| **Nativity** |  |  | <0.001 |
| US (50 states/DC/territories)-born | 88.3 | 92.8 |  |
| Foreign-born | 11.8 | 7.2 |  |
| **Educational Attainment (ages 25+ only)** |  |  | <0.001 |
| 8th grade or lower | 7.0 | 3.5 |  |
| 9-12th grade, no diploma | 23.1 | 16.6 |  |
| High school/GED | 52.1 | 51.3 |  |
| Some college | 11.0 | 14.7 |  |
| Associate/bachelor's degree | 6.1 | 11.9 |  |
| Graduate degree | 0.7 | 2.0 |  |
| **Marital Status (ages 18+ only)** |  |  | <0.001 |
| Single, never married | 60.9 | 55.3 |  |
| Married | 11.5 | 17.8 |  |
| Divorced | 24.6 | 22.9 |  |
| Widowed | 3.0 | 4.0 |  |
| **Occupation in Lifetime (ages 18+ only, years 2022-2024)** |  |  |  |
| Construction/extraction | 15.9 | 15.2 | 0.420 |
| Building/grounds cleaning/maintenance | 3.8 | 4.4 | 0.223 |
| Farming/fishing/forestry | 0.8 | 0.8 | 0.949 |
| **US Census Division of Residence** |  |  | <0.001 |
| New England | 0.5 | 5.7 |  |
| Middle Atlantic | 1.5 | 13.9 |  |
| East North Central | 2.6 | 15.7 |  |
| West North Central | 1.6 | 4.8 |  |
| South Atlantic | 3.1 | 22.1 |  |
| East South Central | 1.8 | 7.3 |  |
| West South Central | 10.3 | 8.7 |  |
| Mountain | 67.8 | 7.5 |  |
| Pacific | 10.9 | 14.3 |  |
| **Overdose/death Circumstances** |  |  |  |
| **Drugs Involved** |  |  |  |
| Synthetic opioids (e.g., fentanyl) | 29.3 | 62.0 | <0.001 |
| Psychostimulants (e.g., methamphetamine) | 85.9 | 28.9 | <0.001 |
| Cocaine | 7.0 | 24.5 | <0.001 |
| Psychostimulants + synthetic opioids | 23.9 | 16.0 | <0.001 |
| Cocaine + synthetic opioids | 1.6 | 17.3 | <0.001 |
| **Injury Occurred at Work (ages 18+ only)** | 1.2 | 0.5 | <0.001 |
| **Place of Injury: at home** | 15.8 | 64.1 | <0.001 |
| **Place of Injury: on the street or highway** | 18.9 | 2.2 | <0.001 |
| **Place of Death** |  |  | <0.001 |
| Medical Center | 32.8 | 22.8 |  |
| Home | 8.7 | 50.4 |  |
| Hospice/Nursing/Long-term-care Facility | 0.5 | 0.5 |  |
| Other | 58.1 | 26.4 |  |

*Notes:* Data from the 2018-2024 Multiple Cause of Death files from the National Center for Health Statistics. “Heat recorded” refers to the presence of an International Classification of Diseases-10 code for X30 or T67 as “multiple cause of death,” without code W92. The “synthetic opioids” category excludes methadone. The “psychostimulants” category excludes cocaine. §P values calculated using chi-square tests for categorical measures and a Wilcoxon rank-sum test for the one continuous measure.
*Abbreviations:* NH, Non-Hispanic; Am. Indian/AK Native, American Indian or Alaska Native.

With respect to circumstances of the overdose and death, the injury (overdose) was reported to have occurred at work in 1.2% of the deaths with, and 0.5% of the deaths without, heat involvement recorded. The proportion of overdoses that had occurred on the street or highway was higher in overdoses with, versus without, heat involvement reported. The proportion that had occurred in a home was relatively lower in deaths with, versus without, heat involvement recorded. Similarly, “home” was the place of death for half of all overdoses without heat involvement reported but only 8.7% of overdoses with heat involvement reported. Psychostimulants, such as methamphetamine, were reportedly involved in 85.9% of overdose deaths with, versus 28.9% of those without, heat involvement, whereas cocaine was involved in a lower share of deaths with (7.0%), versus without (24.5%), heat recorded. Synthetic opioids, such as fentanyl, were involved in 29.3% of overdose deaths with, versus 62.0% of deaths without, heat recorded. In the supplemental analysis excluding Arizona resident deaths (supplemental Table S2), overall patterns were relatively similar to the main results presented in Table 1.

## Discussion

In this analysis of death certificate data from all US resident deaths, overdose deaths with recorded heat involvement rose from fewer than 10 in 2001 to 558 in 2024. These deaths were concentrated in the Southwest, particularly in Arizona and Nevada, differing from overdose deaths without recorded heat involvement in terms of the demographics of those most affected, the drugs most frequently involved, and the locations of overdose and death. The majority (86%) of these deaths involved psychostimulants such as methamphetamine.

In analyses of mortality rates over time, overdose deaths with recorded heat involvement were relatively stable from 2005-2013, yet followed a marked increase from 2013-2024, with an estimated annual percent change of 30%. This pattern does not closely align with the trend in overdose deaths overall, which followed a long-term exponential growth curve beginning in 1979 before declining in 2023-2024 (Friedman et al., 2026). Instead, the observed trend more closely resembles trends in US heat-related mortality rates overall, which began increasing significantly in 2016 (Howard et al., 2024), and trends in overdose deaths involving psychostimulants, which began escalating after 2010 (Garnett & Miniño, 2024). Nonetheless, whereas psychostimulant-involved overdose deaths in the US plateaued after 2021 (Garnett & Miniño, 2024) and declined from 2023-2024 (Garnett & Miniño, 2026), overdose deaths with recorded heat involvement did not decline during the study period. The continued increase in overdose deaths with heat documented may reflect the increasing frequency of extreme heat events (Environmental Protection Agency, 2024) and social contexts that elevate risk of heat-related illness; at the same time, the trends in the present study rely on what is reported on death certificates. Prior research has identified improvements over time in the completeness of drug reporting on death certificates in the US (Gutkind et al., 2025). It is therefore plausible that improvements in death investigation practices (as well as in the identification, documentation, and coding of multiple causes of death) contributed to the trend observed in this study, as the extent of underreporting for overdose deaths with heat involvement may have lessened over time.

Overdose deaths with recorded heat involvement were concentrated in residents of Arizona and Nevada, which also exhibited the highest mortality rates and percentages of overdose deaths with heat involvement recorded. Both Arizona and Nevada are home to urban areas with large populations (e.g., the Phoenix and Las Vegas metropolitan areas) that experience extreme heat. Nonetheless, variation across states should be interpreted with caution, as cause-of-death reporting practices may differ between states and between jurisdictions that use medical examiners versus coroners for their medicolegal death investigation system. Such jurisdictional differences have been shown to influence completeness of reporting drug involvement on death certificates (Gutkind et al., 2025) and may also affect the identification and documentation of other cause of death reporting, including heat involvement.

With respect to patterns observed in overdose circumstances, home was the recorded place of injury (overdose) for only 16% of overdose deaths with, versus 64% of those without, heat involvement recorded. Moreover, nearly one in five (19%) overdose deaths with heat involvement recorded reportedly occurred on the street or highway. These results might be partially attributed to the observation that exposure to extreme heat is generally more intense outdoors than inside a home. Although death certificates do not include information about decedents’ living situations, in an analysis of SUDORS data from Arizona, evidence of homelessness was documented in 60% of individuals who died of an overdose with heat involvement recorded (Cano et al., 2025). As such, while the present study was not able to directly assess the involvement of homelessness in the deaths analyzed, prior evidence linking homelessness to heat-related illness (Weckstein et al., 2025) and to heat-related overdose deaths (Cano et al., 2025) is consistent with several of the study’s findings, including patterns in place of injury and place of death.

When examining demographic patterns, Mexican-heritage, American Indian/Alaska Native, and foreign-born populations were overrepresented in overdose deaths with (versus without) heat recorded. One potential explanation is that Mexican-heritage and American Indian populations are more concentrated (Moslimani et al., 2023; Sánchez-Rivera et al., 2023) in states with the most extreme heat, including states that account for a large portion of all overdose deaths with heat involvement recorded in the US. At the same time, because homelessness is a notable risk factor for heat involvement in overdose deaths (Cano et al., 2025), racial disparities in homelessness (Cano et al., 2024; De Sousa & Henry, 2024) may also influence these patterns. The finding that foreign-born decedents accounted for a larger share of the overdose deaths with (versus without) recorded heat involvement is reminiscent of a prior study documenting a higher risk of heat-related mortality for non-citizens, compared to citizens, in the US (Taylor et al., 2018). One possible contributing factor is that immigrants in the US are disproportionately represented in occupations that require outdoor labor, such as agriculture, construction, or landscaping (Moyce & Schenker, 2017). In the present study, only 1.2% of the overdoses with heat involvement recorded had reportedly occurred at work; at the same time, this figure may not include all cases involving heat exposure that began at work, as the physiological effects of heat exposure during the workday may persist even after an employee leaves work for the day (State of California Department of Industrial Relations, 2011). Consistent with a recent analysis of Arizona data (Cano et al., 2025), which found methamphetamine present in 92% of overdose deaths with documented heat involvement, in the present study, psychostimulants (e.g., methamphetamine) were involved in 85.9% of overdose deaths with heat involvement reported. Suggesting a distinct drug-environment context, overdose deaths with heat involvement were also disproportionately concentrated in the Mountain region (67.8%), particularly in states such as Arizona and Nevada, which are characterized by extreme summer heat, and were far less likely to occur at home but more likely to occur on streets or highways. Neurobiologically, psychostimulants amplify heat-related risk through increased metabolic heat production, disruption of central thermoregulatory processes, and vasoconstriction that reduces heat dissipation (Matsumoto et al., 2014; Callaway & Clark, 1994). In the presence of environmental heat exposure, these effects can produce potentially lethal hyperthermia. The study findings also point to prior studies that have documented a relationship between methamphetamine use and homelessness (Daniulaityte et al., 2020; Mehtani et al., 2023), as individuals may rely on methamphetamine as a survival strategy to help manage hardships and vulnerabilities related to homelessness (Al-Tayyib et al., 2017; Damon et al 2019). Preliminary qualitative data from Arizona suggest that although many people who use drugs recognize methamphetamine risks in terms of potential dehydration and “overheating,” they also indicated that they relied on methamphetamine as a tool to help tolerate the misery and challenges of extreme heat, especially when experiencing housing instability (Daniulaityte et al., 2025).

Despite evidence that cocaine can also induce hyperthermia, including fatal hyperthermia (Callaway & Clark, 1994; Crandall, Vongpatanasin, & Victor, 2002; Bauwens, Boggs, & Hartwell, 1989), we found that cocaine was notably underrepresented (7.0%) in overdose deaths with heat involvement recorded. The underrepresentation of cocaine in overdose deaths with heat involvement recorded may reflect geographical patterns of drug use; compared to the eastern US, there is a notably lower prevalence of cocaine involvement in overdose deaths in general in the western US, where overdose deaths with recorded heat involvement are concentrated (CDC, 2026). At the same time, methamphetamine’s longer half-life (approximately 10 hours; Cruickshank & Dyer, 2009) may plausibly result in prolonged thermoregulatory dysfunction, whereas cocaine’s shorter half-life (1-1.5 hours; Benowitz, 1993) produces more acute effects that may be less likely to result in sustained hyperthermia. Overall, the study findings suggest that prolonged stimulant effects, environmental heat exposure, and specific settings may all interact to increase the likelihood of a fatality.

## Limitations

As an analysis of mortality records, this study is subject to several limitations of death certificate reporting. Although reporting has improved over time, the completeness of death certificate reporting of specific drugs involved in overdose deaths varies between jurisdictions (Gutkind et al., 2025). Similarly, as noted previously, it is likely that the extent to which heat-involvement (when present) would be identified, reported, and coded on a death certificate may differ over time and between jurisdictions. Because the dataset used in this study only included ICD-10 codes for causes of death, we were unable to analyze literal text from death certificates to identify additional relevant deaths not captured in the ICD-10 codes listed. As a result, under-identification, underreporting, or under-coding of heat involvement in deaths may impact estimates of temporal trends, state variation, and characteristics of deaths. Overall, results of this study reflect patterns observed in overdose deaths with heat involvement specifically recorded on the death certificate and may not generalize to heat-related overdose deaths that were not coded accordingly on the death certificate. In addition, key variables of interest, such as education, race/ethnicity, occupation, and place of injury are sometimes missing on death certificates, and certain racial/ethnic categories such as NH AI/AN are underreported (Arias et al., 2016). The mortality data used in this study also lacked several relevant measures of interest, such as the decedent’s living conditions (e.g., homelessness), drug use patterns, or health. Finally, broader environmental exposures, such as weather conditions and ambient temperature, were outside the scope of this analysis of individual-level data.

## Data Availability

Data used in the study are restricted-access through the U.S. National Center for Health Statistics

---

**Supplemental Table S1.** Levels of missingness for each measure used in the study.

| Measure | % Missing in overall sample | % Missing in overdose deaths with heat involvement recorded | % Missing in overdose deaths without heat involvement recorded | Notes |
| --- | --- | --- | --- | --- |
| **Year of Death** | 0 | 0 | 0 |  |
| **Month of Death** | 0 | 0 | 0 |  |
| **State of Residence** | 0 | 0 | 0 |  |
| **Age** | 0.01% | 0.05% | 0.01% |  |
| **Sex** | 0 | 0 | 0 |  |
| **Race/ethnicity** | 0.77% | 2.26% | 0.76% | Missing data were from missing responses regarding whether decedent was of Hispanic origin. |
| **Nativity** | 0 | 0 | 0 |  |
| **Education** (for ages 25+ only) | 3.76% | 14.23% | 3.73% |  |
| **Marital Status** (for ages 18+ only) | 2.52% | 10.55% | 2.49% |  |
| **Usual Occupation in Building/grounds cleaning/ maintenance** (ages 18+ only, years 2022-2024) | * | * | * | *In this measure, any other occupation response (including missing) was coded "no." Only 0.06% of occupation responses were originally missing in the analytic sample. |
| **Usual Occupation in Farming/ fishing/ forestry** (ages 18+ only, years 2022-2024) | * | * | * | *In this measure, any other occupation response (including missing) was coded "no." Only 0.06% of occupation responses were originally missing in the analytic sample. |
| **Usual Occupation in Construction** (ages 18+ only, years 2022-2024) | * | * | * | *In this measure, any other occupation response (including missing) was coded "no." Only 0.06% of occupation responses were originally missing in the analytic sample. |
| **US Census Division of Residence** | 0 | 0 | 0 |  |
| **Drugs Involved** | 0 | 0 | 0 | Any drug category was coded "yes" if the corresponding ICD-10 code was included as a cause of death, and "no" otherwise. The absence of a drug reported on a death certificate does not necessarily mean this drug was not involved or detected, but only that it was not recorded. |
| **Injury Occurred at Work** (ages 18+) | 6.05% | 2.54% | 6.06% |  |
| **Place of Injury- at Home** | * | * | * | *In this measure, any response other than "home" was coded as "no." However, the category "unspecified place" was reported in 19.32% of the sample, and one additional response was missing. |
| **Place of Injury- on a Street/Highway** | * | * | * | *In this measure, any response other than "home" was coded as "no." However, the category "unspecified place" was reported in 19.32% of the sample, and one additional response was missing. |
| **Place of Death** | 0.04% | 0.00% | 0.04% |  |

**Supplemental Table S2.** State-level data (counts, percentages, and mortality rates) for drug overdose deaths with heat involvement recorded on the death certificate, 2001-2024.

| State | Number of Overdose Deaths with Heat Recorded | Mortality Rate per 100,000 (Overdose Deaths with Heat Recorded) | Percent of Overdose Deaths with Heat also Recorded |
| --- | --- | --- | --- |
| Alabama | 10 | 0.009 | 0.06 |
| Alaska | * |  |  |
| Arizona | 1,185 | 0.752 | 3.53 |
| Arkansas | 25 | 0.036 | 0.28 |
| California | 258 | 0.029 | 0.20 |
| Colorado | 19 | 0.015 | 0.08 |
| Connecticut | * |  |  |
| Delaware | * |  |  |
| District of Columbia | * |  |  |
| Florida | 40 | 0.008 | 0.04 |
| Georgia | 12 | 0.005 | 0.04 |
| Hawaii | * |  |  |
| Idaho | * |  |  |
| Illinois | 24 | 0.008 | 0.05 |
| Indiana | 15 | 0.010 | 0.05 |
| Iowa | * |  |  |
| Kansas | 15 | 0.022 | 0.18 |
| Kentucky | 18 | 0.017 | 0.06 |
| Louisiana | 44 | 0.040 | 0.18 |
| Maine | * |  |  |
| Maryland | 21 | 0.015 | 0.06 |
| Massachusetts | * |  |  |
| Michigan | * |  |  |
| Minnesota | * |  |  |
| Mississippi | * |  |  |
| Missouri | 25 | 0.017 | 0.09 |
| Montana | * |  |  |
| Nebraska | * |  |  |
| Nevada | 436 | 0.656 | 2.84 |
| New Hampshire | * |  |  |
| New Jersey | * |  |  |
| New Mexico | 26 | 0.053 | 0.20 |
| New York | 19 | 0.004 | 0.03 |
| North Carolina | 10 | 0.004 | 0.02 |
| North Dakota | * |  |  |
| Ohio | 26 | 0.009 | 0.04 |
| Oklahoma | 32 | 0.035 | 0.19 |
| Oregon | 19 | 0.020 | 0.12 |
| Pennsylvania | 28 | 0.009 | 0.04 |
| Rhode Island | * |  |  |
| South Carolina | * |  |  |
| South Dakota | * |  |  |
| Tennessee | 21 | 0.014 | 0.05 |
| Texas | 180 | 0.029 | 0.26 |
| Utah | 13 | 0.019 | 0.10 |
| Vermont | * |  |  |
| Virginia | * |  |  |
| Washington | 38 | 0.023 | 0.12 |
| West Virginia | * |  |  |
| Wisconsin | * |  |  |
| Wyoming | * |  |  |

*Note.* \*Due to confidentiality regulations, numbers not presented for any state with fewer than 10 deaths over the period examined (2001-2024).

**Supplemental Table S3.**
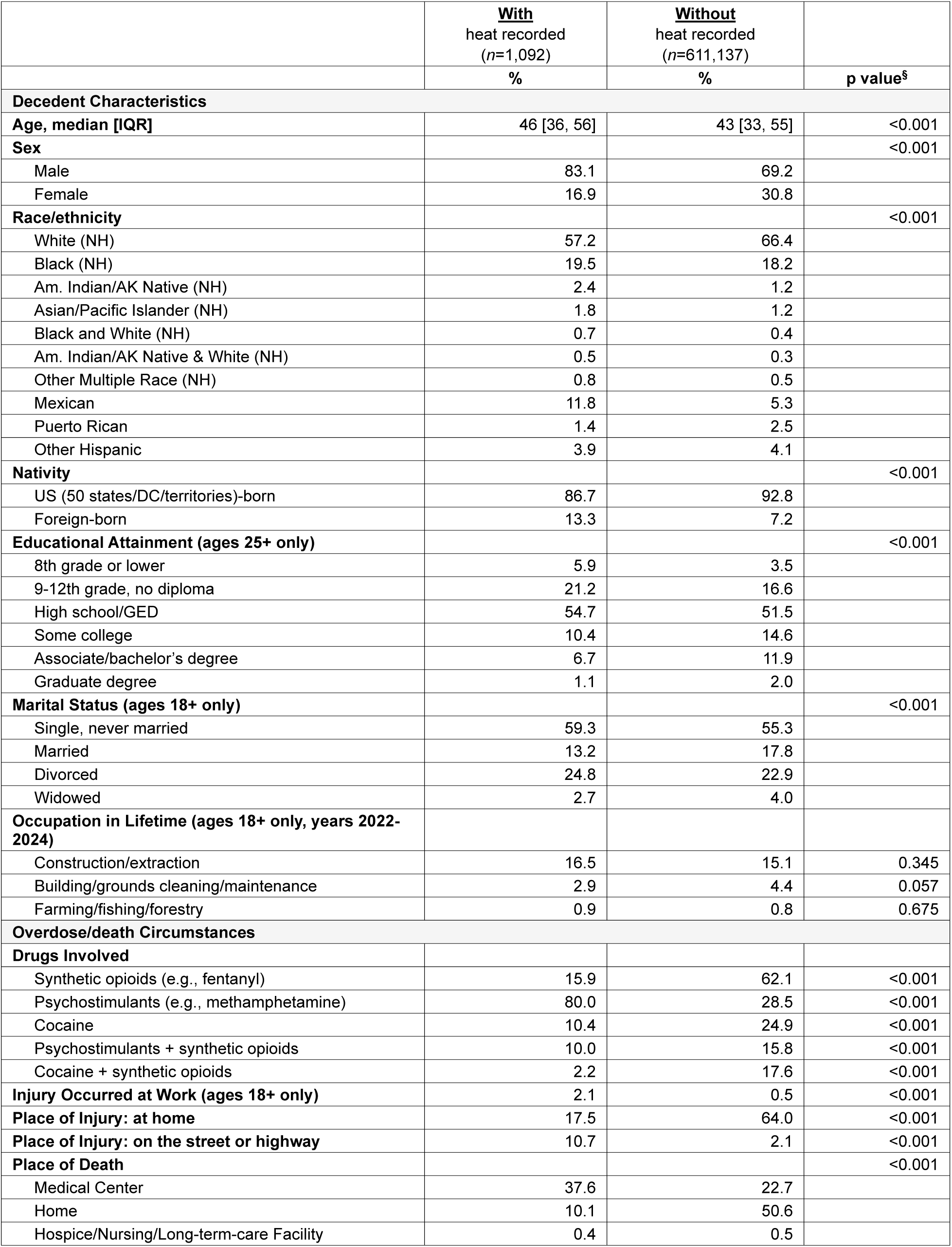
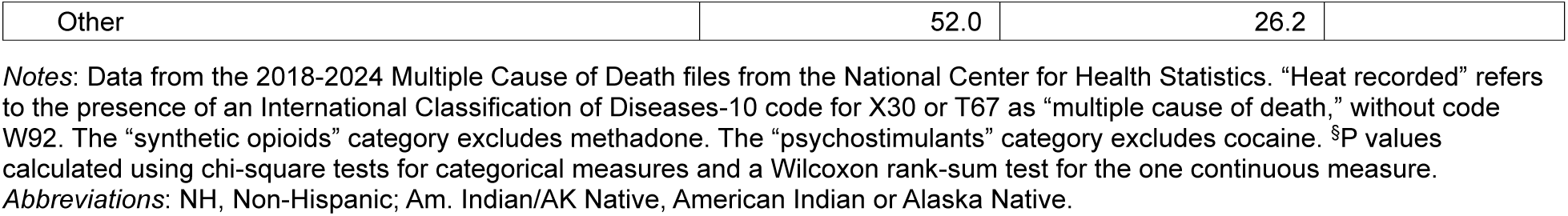
Characteristics of overdose deaths with, versus without, heat involvement recorded, United States, 2018-2024, <u>excluding Arizona resident deaths</u>.

